# Tumors with mutations in chromatin regulators are associated with higher mutational burden and improved response to checkpoint immunotherapy

**DOI:** 10.1101/2024.10.15.24315153

**Authors:** Goran Kungulovski, Marija Gjorgjievska, Sanja Mehandziska, Djansel Bukovec, Milan Risteski, Ivan Kungulovski, Zan Mitrev

**Affiliations:** Zan Mitrev Clinic, Skopje, Republic of Macedonia; Fingerprint Diagnostics LLC, Skopje, Republic of Macedonia

**Author notes:** Corresponding author: Dr. Goran Kungulovski, Fingerprint Diagnostics LLC Ivan Agovski 7-1, 1000, Skopje, Republic of Macedonia.

**Keywords:** cancer epigenetics, cancer genetics, chromatin, immunotherapy, checkpoint inhibitors, tumor mutational burden

## Abstract

In recent years, it has been demonstrated that many of the pervasive genetic defects throughout cancerogenesis occur in genes encoding chromatin regulators (CRs). We analyzed the distribution and characteristics of well-studied CRs across tens of thousands of tumor samples. Our analysis revealed that tumors with mutations in CRs are associated with high tumor mutational burden (TMB). The co-occurrence of mutations in multiple CRs was linked with a further increase in TMB. Given that TMB may predict the clinical response to immune checkpoint inhibitor (ICI) treatment, we investigated the relationship between mutations in CRs and ICI response. We found that patients harboring mutations in CRs exhibited improved responses to ICI treatment, comparable to those with deficiencies in canonical DNA repair pathways. Overall, this study uncovered significant relationships between mutations in chromatin regulators and critical features of cancer, underscoring the need for further functional and clinical studies.

## INTRODUCTION

Biologically, cancer denotes a multifaceted group of diseases, typically characterized by cell regulation failure, which equips the cells with the potential to divide in an uncontrolled manner, evade the immune system, and over time, invade and spread into neighboring tissues and organs of the body (Hanahan, 2022). Traditionally, cancer has been thought to be driven by the accumulation of genetic defects such as mutations, deletions, amplifications, and translocations in so-called oncogenes and tumor-suppressor genes (Kontomanolis et al., 2020). Consequently, most of the endeavors undertaken in cancer biology research have historically focused on genetic aberrations in genes with prominent roles in DNA repair, cell proliferation, and cell signaling.

However, in recent years, the widespread application of next-generation sequencing (NGS) technologies for comprehensive cancer profiling has revealed that many of the pervasive genetic defects during carcinogenesis occur in chromatin (epigenetic) regulators (CRs) (Rodrigues-Paredes & Esteller, 2011; Dawson & Kouzarides, 2012; Yu et al., 2024). Such mutations in chromatin writers, readers, erasers, remodelers, and even the histone proteins themselves, can rewire the chromatin landscape of cells, leading to altered epigenetic states and expression programs that contribute to the biological and clinical aspects of tumorigenesis (Baylin & Jones, 2016; Timp & Feinberg, 2013; Feinberg et al., 2016). Hence, a deeper understanding of the distribution, interrelationships, and features of chromatin regulators in cancer can significantly aid our understanding of cancer biology, and treatment.

In this study, we sought to examine the features of a subset of mutated chromatin regulators across tens of thousands of available genetic maps from various cancer types. We uncovered a strong positive relationship between the presence of mutations in CRs and TMB, which in turn was associated with significantly improved survival following treatment with ICI.

## RESULTS

### Mutations in chromatin regulators are pervasive in cancer and associated with high tumor mutational burden

We commenced by examining the distribution and characteristics of mutations in well-studied chromatin regulators (Timp & Feinberg, 2013; Feinberg et al., 2016) **(Suppl. Data 1)** across tens of thousands of tumor samples. This list is by no means exhaustive in capturing all chromatin regulators frequently mutated in cancer. However, it includes many well-studied CRs that are commonly captured by traditional molecular profiling panels and described in (Timp & Feinberg, 2013; Feinberg et al., 2016). We utilized data from large studies such as the TCGA, MSK-ICI, MSK-MET, PCAWG, and the OrigiMed deposited in the cBioportal platform (TCGA Research Network, 2013; Zehir et al., 2017: Samstein et al., 2019; Consortium, 2020; Nguyen et al., 2022; Wu et al., 2022). Our analysis revealed that mutations in chromatin regulators were pervasive in cancer, especially enriched in particular cancer types such as endometrial, lung, esophagogastric, colorectal, melanoma, and bladder cancer **(Suppl. Figures 1, and 2)**. Given that many of these cancer types are known to be associated with a high mutational load **(Suppl. Figure 2C)** (Chalmers et al., 2017), we sought to ascertain the relationship between the presence of mutations in chromatin regulators and TMB. Tumors with high TMB will theoretically correlate with the probability of a somatic mutation in any part of the genome, including in chromatin regulators (CRs). To demonstrate that the prevalence and distribution of CR mutations are distinct and non-random in high-TMB tumors, we plotted the enrichment of 100 randomly selected non-CR genes alongside canonical tumor suppressors and oncogenes and indeed observed distinct patterns **(Suppl. Figure 2A and B).** Tumors with mutations in the selected CRs had a higher TMB compared to the general cohort or tumors without these CR mutations (**Figure 1A and B, p <0.0001 Kruskal-Wallis test**). Then we wanted to investigate the relationship between tumors harboring single CRs and their effect on TMB levels. We calculated the TMB in tumor samples harboring at least one mutation in a CR gene, in tumors harboring mutations in canonical oncogenes or tumor suppressors, or in tumors harboring mutations in in canonical DNA repair pathways such as the mismatch repair (MMR) machinery or POLE/POLD1 as controls. The presence of mutations in the MMR or POLE/POLD1 machinery has been reported as a proxy marker for high TMB, and ICI-response (Le et al., 2015; Ma et al., 2022). Our results demonstrated significantly higher TMB levels in tumors harboring mutations in certain CRs (e.g *SMARCD1, NSD2, DNMT1, BRD4, TET2, TET1, DNMT3B, EZH1/2*) compared to MMR or POLE/D1 mutated tumors (p < 0.0001, Dunnett’s multiple comparisons test) (**Figure 1C and D; Suppl. Figures 3 and 4; Suppl. Data 2-5**). The types of CRs associated with high TMB were reproducible across datasets, indicating the non-random distribution of CRs in tumor samples (**Suppl. Figure 5**).

**Figure 1.**
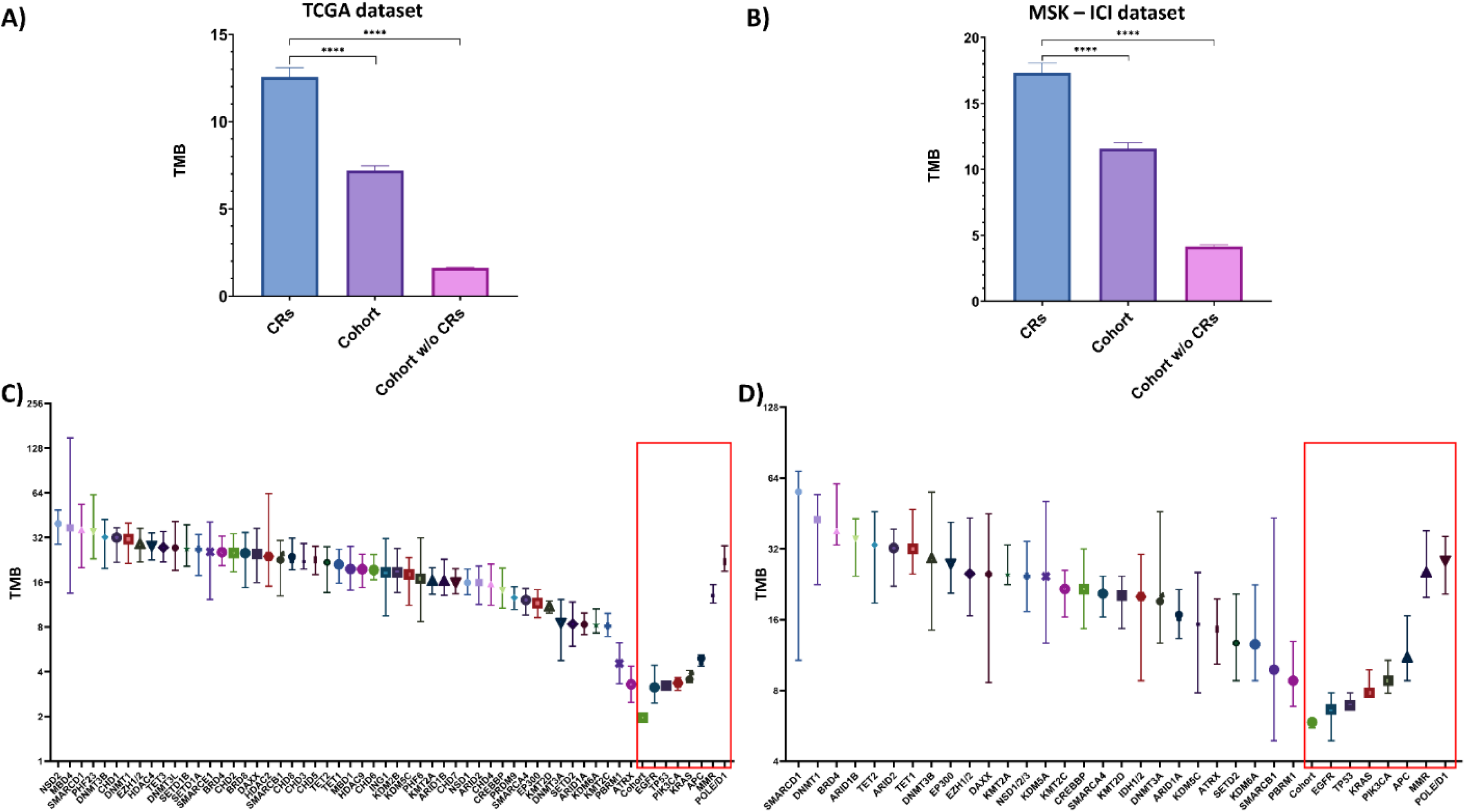
Distribution of tumor mutational burden (TMB) in relevant groups of samples. A) Samples with mutations in chromatin regulators (CRs) n=5321, the cohort n=10443, and the cohort with CR genes removed n = 5122, taken from the TCGA dataset, B) Samples with mutations in chromatin regulators (CRs) n=935, the cohort n=1661, and the cohort with CR genes removed n=726, taken from the MSK-ICI dataset. The bars show the mean of the group, while the error bars represent the standard error of the mean (SEM). Statistical significance was calculated with the Kruskal-Wallis test. C) Samples with mutations in single chromatin regulators (CRs), samples with mutations in canonical oncogenes and tumor-suppressor genes, and samples with mutations in MMR-genes and *POLE*, and *POLD1*, taken from the TCGA dataset. The median of the analyzed samples is 312. D) Samples with mutations in single chromatin regulators (CRs), samples with mutations in canonical oncogenes and tumor-suppressor genes, and samples with mutations in MMR-genes and *POLE* and *POLD1*, taken from the MSK-ICI dataset. The median of the analyzed samples is 102. The graph represents the median with 95% confidence intervals (CI). The red boxes indicate the positive and negative controls. Supplementary Figures 1-5 are related to this figure. The statistical significance between samples can be found in Supplementary Data 2-5 calculated with the Dunn’s multiple comparisons test. The number of samples per subgroup used in this study can be found in Suppl. Data 9

### The co-occurrence of mutations in chromatin regulators leads to additional increases in TMB

Next, we were curious to ascertain if the co-occurrence of mutations in two or more CRs would have an enhancing effect on the levels of TMB as displayed in the red box in (**Figure 2A**). Indeed, by analyzing the MSK-ICI data, we observed that the simultaneous presence of mutations in multiple CRs had a strong linear relationship with TMB levels (Pearson’s r = 0.87, R^2^ = 0.76, p < 0.0001) (**Figure 2B**). A positive relationship was also observed when mutations were simultaneously present in multiple genes of the MMR machinery and/or POLE/D1 (Pearson’s r =0.68), albeit with lower predictability (R^2^ = 0.46) (**Suppl. Figure 6A**). This was not the case when TMB levels were assessed in association with co-occurring mutations in canonical oncogenes and tumor suppressors (Pearson’s r = 0.43, R^2^ = 0.18) (**Suppl. Figure 6B**). Then we selected the top 10 high-TMB CRs from the MSK-ICI and TCGA datasets (**Figure 2A; Suppl. Figure 1**). By calculating the log2 odds ratio against all other CRs, and controls (MMR and POLE/D1) we discovered statistically significant associations of co-occurrence (**Suppl. Data 6 and 7**), which again led to significant increases of TMB (p < 0.0001, Dunnett’s multiple comparisons test) (**Figure 2C; Suppl. Figures 7, and 8**). A similar cumulative effect leading to increased levels of TMB was observed when mutations in CRs and MMR or POLE/D1 were present at the same time, with CRs having a stronger effect on the TMB levels in MMR and POLE/D1 samples than vice versa (**Suppl. Figure 9A and B**). Similar observations were made when we selected the top 10 high TMB-associated CR and non-CR genes (out of 100 randomly selected non-CR genes), and showed that the top 10 CRs had a stronger effect on the TMB levels in top 10 non-CR samples than vice versa, again showcasing the effect of CRs on TMB **(Suppl. Figure 9C and D)**. All in all, these observations were consistently corroborated across multiple independent datasets, demonstrating that single or co-occurring mutations in chromatin regulators or other high TMB-associated genes are strongly linked to higher TMB.

**Figure 2.**
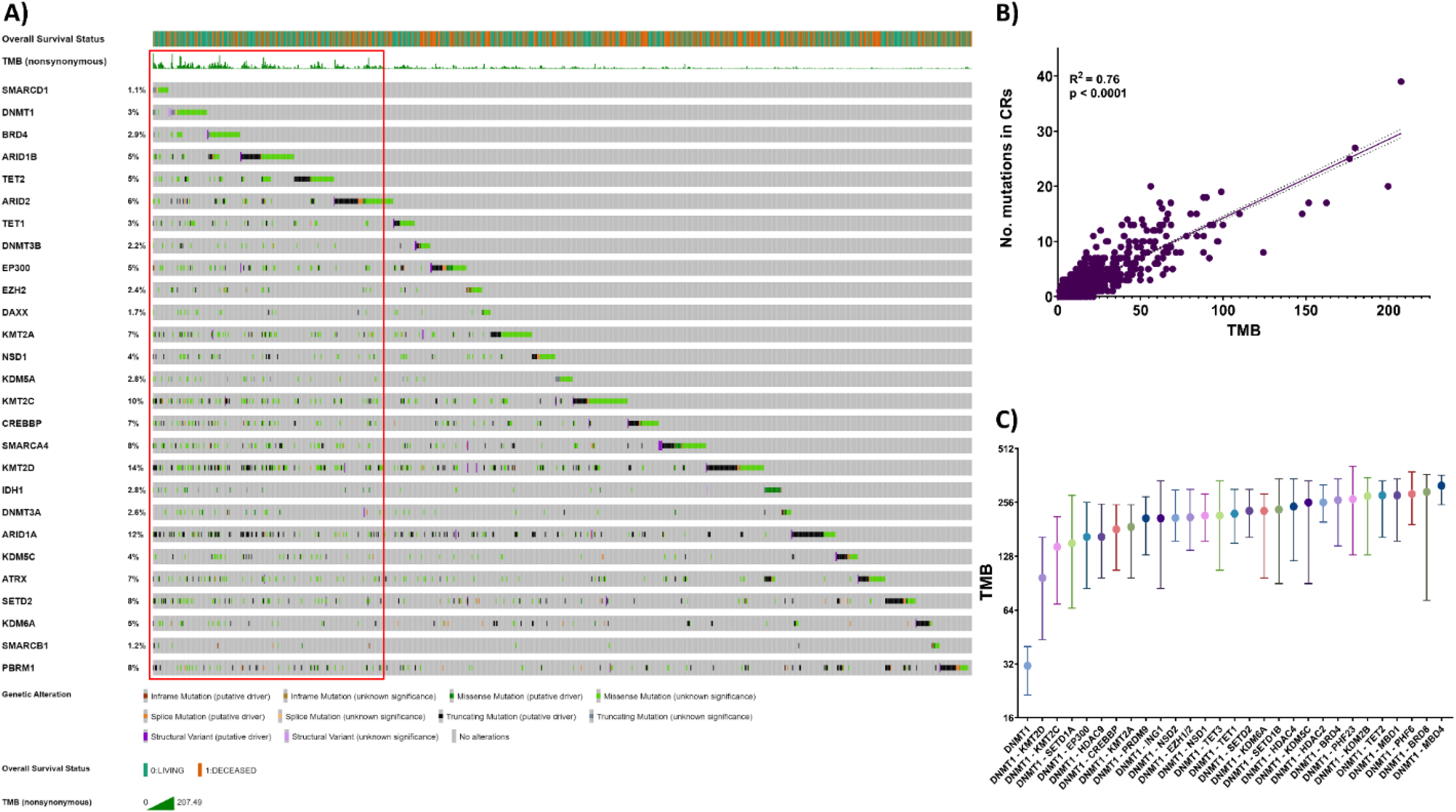
Co-occurrence of chromatin regulators leads to increased TMB. A) Oncoprint analysis of the distribution and prevalence of genomic alterations in CRs corresponding to overall survival status, and TMB. The red box visually displays the association of co-occurring mutations in CRs with increased levels of TMB, B) Simple linear regression model showing that TMB can be predicted based on the number of co-occurring mutations in multiple CRs. The datasets are taken from the MSK-ICI study. C) Statistically significant associations of co-occurrence between DNMT1 with other CRs lead to a further increase in TMB. The graph represents the median with 95% confidence intervals (CI). The datasets are taken from the TCGA study. Supplementary Figures 1, 6-9 are related to this figure. The statistical significance between samples can be found in Supplementary Data 6 and 7. The number of samples per subgroup used in this study can be found in Suppl. Data 9.

### Mutations in chromatin regulators lead to improved response to checkpoint immunotherapy

Next, we set out to examine the association between the presence of mutations in CRs, and overall survival after treatment with ICI. We used the MSK-ICI cohort (Samstein et al., 2019), which included 1661 patients whose tumors were profiled with the MSK-IMPACT assay, and who had received at least one dose of ICI as monotherapy or combination. As indicated in the original study, the overall survival was measured from the date of the first ICI treatment to the time of death or most recent follow-up (median 19 months, range 0-80). We corroborated the initial observation showing that higher TMB levels are associated with better outcomes (**Suppl. Figure 10**). Further on, we defined subgroups based on the presence of mutations in selected CRs (e.g. top 10, high TMB CRs), MMR, or POLE/D1 as positive controls (likely responders) versus the average of the entire cohort. The analysis showed that the group of high TMB CRs was associated with improved survival compared to the entire cohort, and on par with MMR or POLE/D1 deficient tumors (**Figure 3А**). Univariate Cox proportional-hazards regression showed the presence of CR mutations was significantly associated with better survival (all CRs, HR = 0.8520, p = 0.0075; TOP5 CRs, HR = 0.5758, p < 0.0007; TOP10 CRs, HR = 0.6845, p < 0.0001), on par with MMR (HR = 0.6162, p = 0.0025), and POLE/D1 (HR = 0.6576, p = 0.0074) deficient patients. Moreover, we analyzed the distribution of mutated CRs in living and deceased patients by calculating the ratio, of a CR in living and deceased patients for each CR individually. We observed a significant enrichment of high TMB CRs in living vs deceased patients (HR ranging from 0.32 up to 1.38) (**Figure 3B; Suppl. Data 8**). To demonstrate that the beneficial effect is attributed to the ICI treatment, and not to a general benefit obtained from the mere presence of mutations in CRs, we compared the ratio of CRs in living and deceased patients taken from the MSK-ICI study (ICI-treated), to those from the MSK-MET, and TCGA studies (variously treated). We observed a low-to-no correlation between these datasets (MSK-ICI vs MSK-MET, r > 0.36 and MSK-ICI vs TCGA, r < –0.06) (**Suppl. Figure 11A and B**). In addition, there was a negligible or no association between high TMB CRs and improved survival in the MSK-MET and TCGA studies (TOP10 MSK-MET genes HR=0.90, p < 0.0003; TOP10 MSK-ICI genes HR=0.95, p=0.05; TOP10 TCGA genes HR=1.01, p = 0.78; TOP10 MSK-ICI genes, HR=0.99, p=0.87) (**Suppl. Figure 11C and D**). In summary, the above data show a strong association between the presence of mutations in selected CRs and improved survival due to ICI treatment.

**Figure 3.**
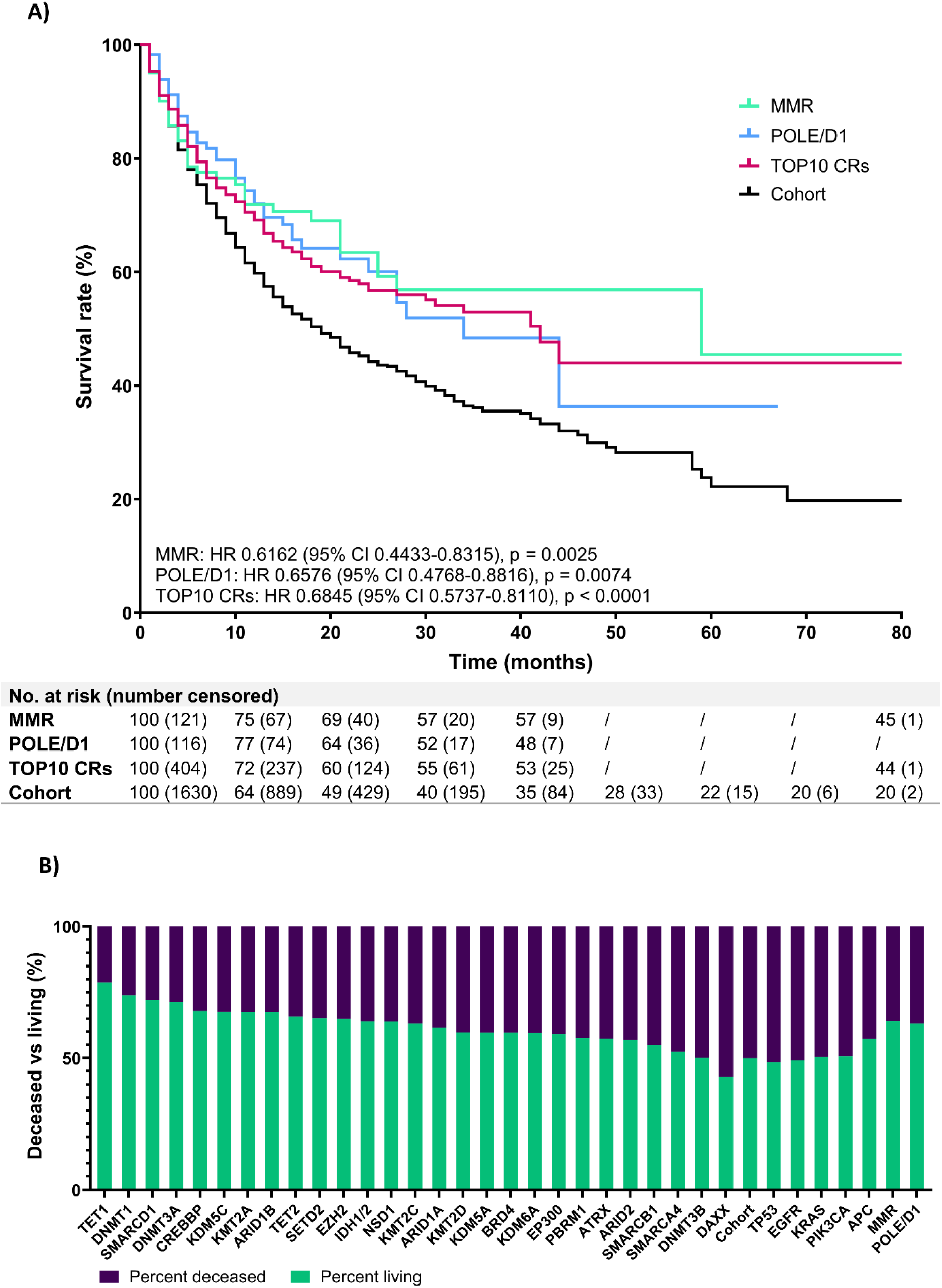
ICI-survival analysis in selected groups of patients taken from the MSK-ICI dataset. A) Kaplan-Meier survival analysis between patients with mutations in the TOP 10 high TMB CRs, mutations in the MMR genes, and mutations in the *POLE* or *POLD1* genes compared with the cohort. The Cox proportional hazards model was used to calculate hazard ratios. B) The distribution of mutated CRs, mutated oncogenes or tumor-suppressors, mutated MMR-genes or *POLE,* and *POLD1* in living and deceased patients who underwent ICI treatment. The datasets are taken from the MSK-ICI study. Supplementary Figures 10 and 11 are related to this figure. The Cox hazard ratios for each CR can be found in Suppl. Data 8. The number of samples per subgroup used in this study can be found in Suppl. Data 9.

## DISCUSSION

The findings of our study highlight the critical role of chromatin regulators in cancer biology and their potential as biomarkers for predicting tumor mutational burden and response to immune checkpoint inhibitors. This is in agreement with previous studies associating CRs such as *SMARCA4* and *ARID1A* with TMB and improve ICI response (Ravi et al., 2023; Wang et al., 2023) Our analysis, which encompassed tens of thousands of tumor samples from publicly available biorepositories, demonstrated that mutations in CRs are widespread across various cancer types and are significantly associated with elevated TMB, particularly pronounced in cancers known to have a high mutational load. This underlines the importance of epigenetic dysregulation in cancer formation, progression, and treatment, and at the same time alludes that some cancers might be more dependent on genetically induced epigenetic changes than others.

Our observation that tumors harboring CR mutations exhibit higher TMB compared to those without such mutations underscores the importance of CRs in influencing the mutational landscape of cancers. Our results indicate that single mutations in CRs are sufficient to increase TMB levels, and this effect is amplified when multiple CR mutations co-occur together or with another high TMB associated gene. The strong linear relationship between the number of CR mutations and TMB, suggests that the detection of mutations CR and repair machinery mutations through the application of small, and cost-effective gene panels could be a reliable predictor of TMB. Although CRs significantly impact TMB, they likely do not fully account for the presence of high TMB in tumors. Further studies are needed to explore the complex relationship between high TMB and other individual genes or gene groups, whether CRs or non-CRs in multivariate analyses.

We also explored the clinical implications of CR mutations in the context of ICI treatment. Despite the heterogeneity, heavy pretreatment, and variable ICI-timing of the MSK-ICI cohort our analysis revealed that patients with tumors harboring CR mutations had improved survival outcomes following ICI therapy. These outcomes were comparable to those with MMR-or POLE/D1 deficient tumors, which are traditionally associated with high TMB and favorable ICI responses. The association of CR mutations with increased levels of TMB, and improved ICI outcomes in such a heterogeneous cohort highlights the robustness of our observations, suggesting that mutations in CRs could serve as clinically meaningful predictive biomarkers to inform clinical decisions regarding ICI treatment, and should be included in tumor profiling panels.

The mechanism by which mutations in CRs systematically dysregulate the epigenome, leading to elevated TMB and improved response can be delineated in two ways. Firstly, the dysregulated epigenome might cause deficiencies in genome maintenance and repair, resulting in a hypermutable state that might yield a higher number of tumor neoantigens, and thus facilitate an improved immune response. Secondly, the dysregulated epigenome might enhance the expressivity of already established neoantigens, facilitating a more robust immune response and better outcomes. Investigating the mechanistic pathways through which CR mutations influence TMB and enhance ICI efficacy could provide deeper insights into cancer immunotherapy and lead to more personalized treatment approaches. Additionally, expanding this research to include other cohorts and cancer types will be crucial to confirm the generalizability of our results. These findings collectively emphasize the need for further functional and clinical studies to validate the role of CR mutations as biomarkers for TMB and ICI response.

In conclusion, our study elucidates the significant relationships between chromatin regulator mutations, tumor mutational burden, and clinical outcomes following ICI treatment. These insights not only advance our understanding of the epigenetic underpinnings of cancer but also pave the way for developing novel biomarkers and therapeutic strategies to improve patient outcomes in cancer immunotherapy.

## MATERIALS AND METHODS

### Datasets used in the study

All genomic datasets used in this study were downloaded from the cBioportal platform, http://www.cbioportal.org/, and were statistically analyzed, and visualized using various tools including Microsoft Excel, Google Sheets, StatsDirect, Prism, and integrated tools within the cBioportal platform, such as Oncoprint (Cerami et al., 2012; Gao et al., 2013). Datasets in the study include the Tumor Cancer Genome Atlas (TCGA; 10967 samples), this is a landmark cancer genomics program, molecularly characterized over 20,000 primary cancer and matched normal samples spanning 33 cancer types; Memorial Sloan Kettering-Metastatis Events and Tropisms (MSK-MET; 25775 samples), this is a pan-cancer cohort of tumor genomic and clinical outcome data and it identifies associations between tumor genomic alterations and patterns of metastatic dissemination across 50 tumor types; Memorial Sloan-Kettering Immune Checkpoint Inhibitors (MSK-ICI; 1661 samples), this is a genomic and survival data from tumor-normal pairs from patients with various cancer types sequenced with the MSK-IMPACT assay; The ICGC/TCGA Pan-Cancer Analysis of Whole Genomes consortium (PCAWG; 2922 samples), this study encompasses whole-cancer genomes and their matched normal tissues across 38 tumor types; and the China PanCancer consortium (OrigiMed; 10194 samples), encompasses the landscape of genomic alterations in solid tumors from the Chinese population. All the datasets are deposited in the cBioportal platform (TCGA Research Network, 2013; Zehir et al., 2017: Samstein et al., 2019; Consortium, 2020; Nguyen et al., 2022; Wu et al., 2022). The number of samples per subgroup used in this study can be found in **Suppl. Data 9**. The TCGA dataset was sequenced using whole exome sequencing (WES), whilst the PCAWG dataset was sequenced with whole genome sequencing (WGS). The remaining datasets were obtained via targeted panel sequencing. Specifically, the MSK-ICI and the MSK-MET datasets were generated using the FDA-approved MSK-IMPACT assay, which targets 341-468 cancer-related genes (Cheng, et al., 2015). The OrigiMed sequences were acquired using the CSYS assay, targeting 450 cancer-related genes (Cao, et al., 2019).

### Statistical analysis

Statistical analyses were conducted using the Kruskal-Wallis non-parametric exact test for multiple-group comparisons to account for distribution differences among the samples. Dunnett’s multiple comparisons test was used to compare each group with a single control. Multiple pairwise comparisons were calculated post-hoc with the Dunn’s test. Survival data were analyzed and interpreted using the Cox proportional hazards model and Kaplan-Meier curves, with comparisons made using the log-rank test. Simple linear regression was employed to model the relationship between TMB and co-occurring mutations. To evaluate the strength of linear relationships between continuous variables, the Pearson correlation coefficient was calculated.

## Declarations

### Ethics approval and consent to participate

Not applicable. All data are taken from publicly available datasets and cancer projects.

### Consent for publication

Not applicable. All data are taken from publicly available datasets and already reported in the literature.

### Availability of data and material

All data are publicly available at the cBioportal repository, http://www.cbioportal.org.

### Author contributions

GK conceived and designed this study. GK, MG, SM, and DjB collected, sorted, curated, and analyzed the data. MR, IK and ZM provided valuable scientific insights. GK wrote the manuscript. All authors contributed to the improvement of the manuscript and read the final version of the manuscript.

## Data Availability

All data are publicly available at the cBioportal repository, http://www.cbioportal.org.

## Acknowledgments

We would like to acknowledge Aleksandar Trifunovski, Aleksandra Horvat, and Bobi Sofronijoski for their assistance in data sorting and integration. We are grateful to Maria Kitanoska and Tamara Cvetkovska for their help in data collection and curation.

