## Supplementary Figures for "Tumors with mutations in chromatin regulators are associated with higher mutational burden and improved response to checkpoint immunotherapy"

**Corresponding author*

### Supplementary Figures

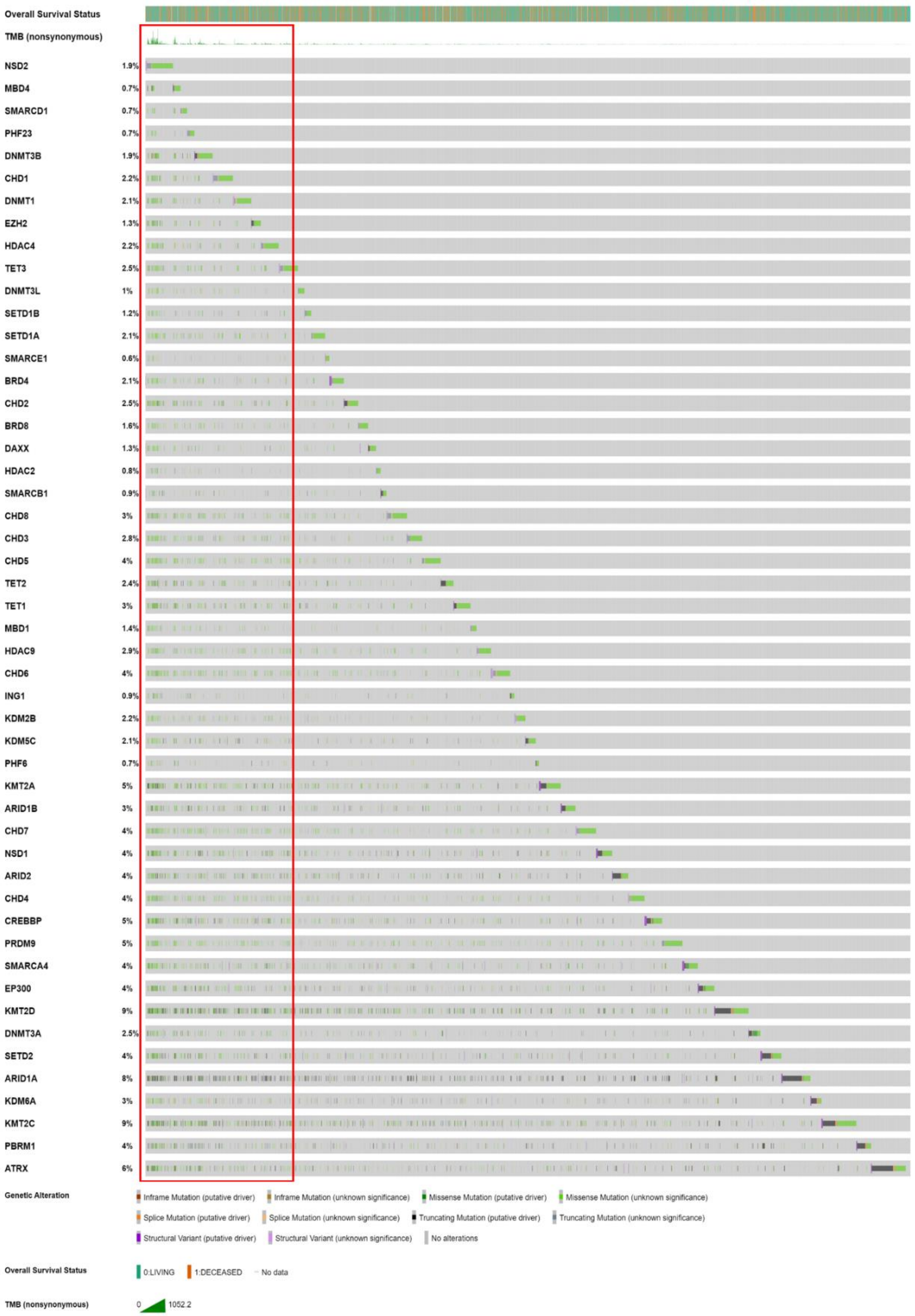
Overall Survival Status
TMB (nonsynonymous)
NSD2 1.9%
MBD4 0.7%
SMARCD1 0.7%
PHF23 0.7%
DNMT3B 1.9%
CHD1 2.2%
DNMT1 2.1%
EZH2 1.3%
HDAC4 2.2%
TET3 2.5%
DNMT3L 1%
SETD1B 1.2%
SETD1A 2.1%
SMARCE1 0.6%
BRD4 2.1%
CHD2 2.5%
BRD8 1.6%
DAXX 1.3%
HDAC2 0.8%
SMARCB1 0.9%
CHD8 3%
CHD3 2.8%
CHD5 4%
TET2 2.4%
TET1 3%
MBD1 1.4%
HDAC9 2.9%
CHD6 4%
ING1 0.9%
KDM2B 2.2%
KDM5C 2.1%
PHF6 0.7%
KMT2A 5%
ARID1B 3%
CHD7 4%
NSD1 4%
ARID2 4%
CHD4 4%
CREBBP 5%
PRDM9 5%
SMARCA4 4%
EP300 4%
KMT2D 9%
DNMT3A 2.5%
SETD2 4%
ARID1A 8%
KDM6A 3%
KMT2C 9%
PBRM1 4%
ATRX 6%
Genetic Alteration
Inframe Mutation (putative driver)
Inframe Mutation (unknown significance)
Missense Mutation (putative driver)
Missense Mutation (unknown significance)
Splice Mutation (putative driver)
Splice Mutation (unknown significance)
Truncating Mutation (putative driver)
Truncating Mutation (unknown significance)
Structural Variant (putative driver)
Structural Variant (unknown significance)
No alterations
Overall Survival Status
0:LIVING
1:DECEASED
No data
TMB (nonsynonymous)
0
1052.2

**Supplementary Figure 1**. Oncoprint analysis of the distribution and prevalence of genomic alterations in CRs corresponding to overall survival status, and TMB. The red box visually indicates the association of co-occurring mutations in CRs with increased levels of TMB. The datasets are taken from the TCGA study. The figure is related to all sections in the main paper.

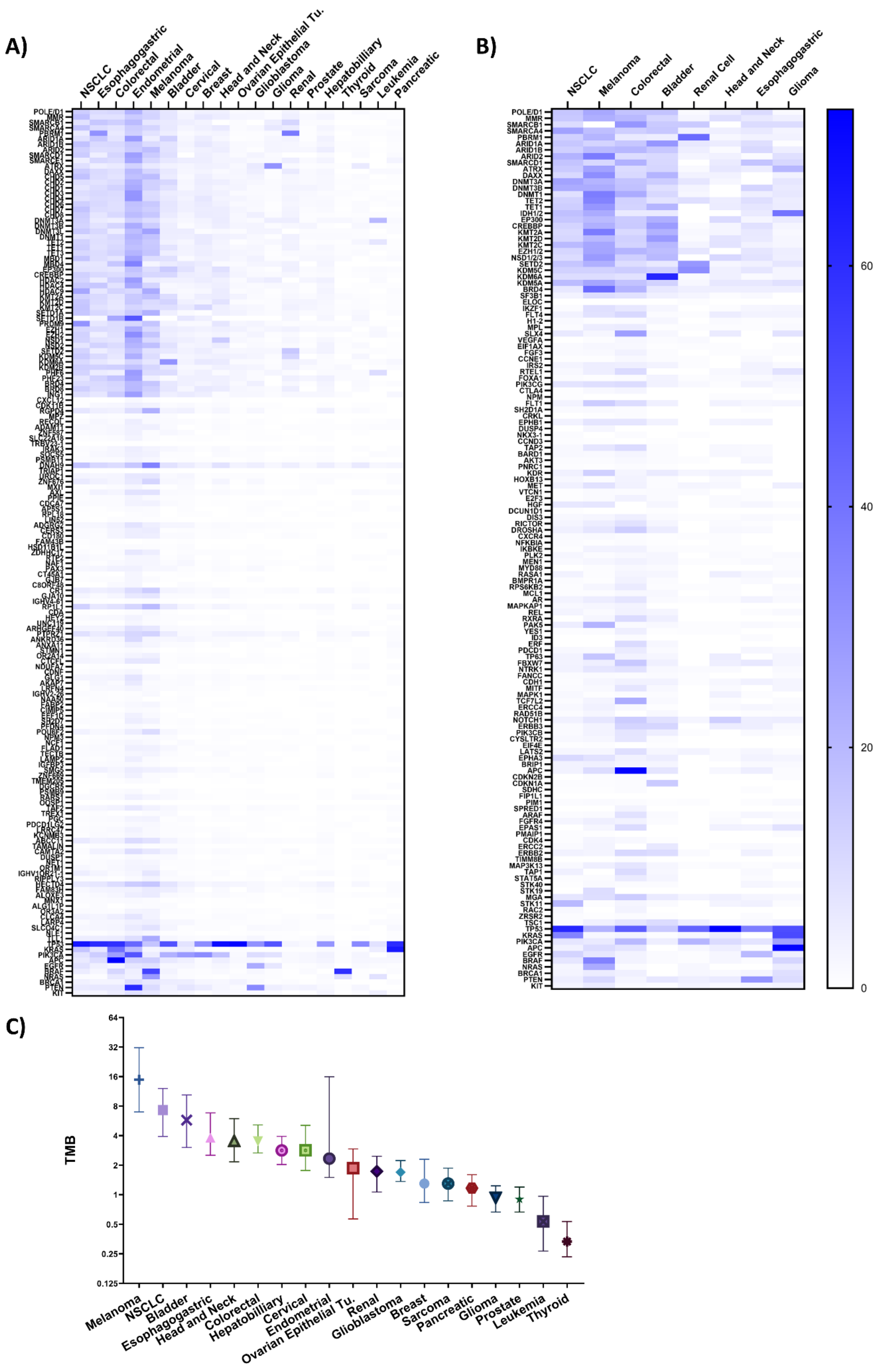
A)
NSCLC
Esophagogastric
Colorectal
Endometrial
Melanoma
Bladder
Cervical
Breast
Head and Neck
Ovarian Epithelial Tu.
Glioblastoma
Glioma
Renal
Prostate
Hepatobilliary
Thyroid
Sarcoma
Leukemia
Pancreatic
B)
NSCLC
Melanoma
Colorectal
Bladder
Renal Cell
Head and Neck
Esophagogastric
Glioma
60
40
20
0
C)
TMB
64
32
16
8
4
2
1
0.5
0.25
0.125
Melanoma
NSCLC
Bladder
Esophagogastric
Head and Neck
Colorectal
Hepatobilliary
Cervical
Endometrial
Ovarian Epithelial Tu.
Renal
Glioblastoma
Breast
Sarcoma
Pancreatic
Glioma
Prostate
Leukemia
Thyroid

**Supplementary Figure 2**. Distribution of mutations in chromatin regulators, 100-randomly selected non-CRs, selection of traditional oncogenes and tumor-suppressor genes and TMB per cancer type. Percentage of enrichment of genes per cancer type; heatmaps represent the datasets from A) the TCGA study, and B) the MSK-ICI study. C) Distribution of TMB per cancer type in the TCGA study. The graph represents the median with 95% confidence intervals (CI). This figure is related to Figure 1. This figure is related to the section "Mutations in chromatin regulators are pervasive in cancer and associated with high tumor mutational burden."

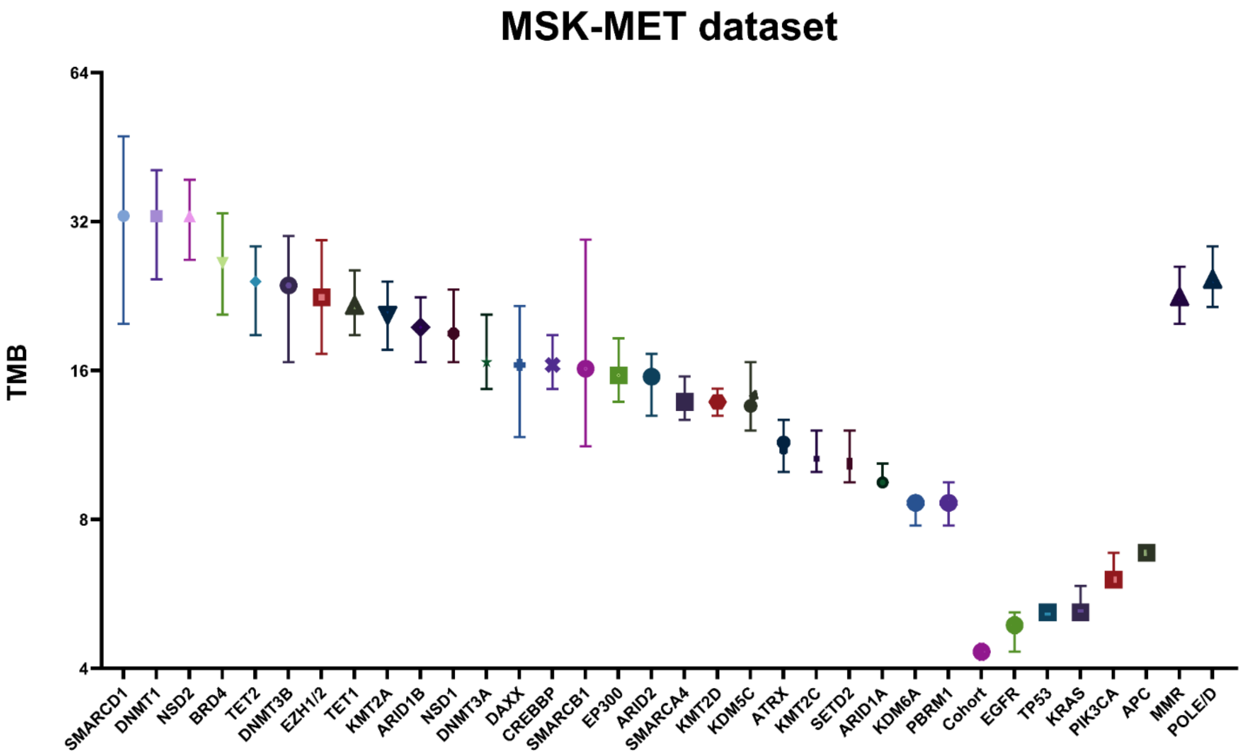


**Supplementary Figure 3**. Samples with mutations in single chromatin regulators (CRs), samples with mutations in canonical oncogenes and tumor-suppressor genes, and samples with mutations in MMR genes and *POLE* and *POLD1* were taken from the MSK-MET dataset. The graph represents the median with 95% confidence intervals (CI). This figure is related to Figure 1. This figure is related to the section "Mutations in chromatin regulators are pervasive in cancer and associated with high tumor mutational burden". The median of the analyzed samples is 916. The number of samples per subgroup used in this study can be found in Suppl. Data 9.

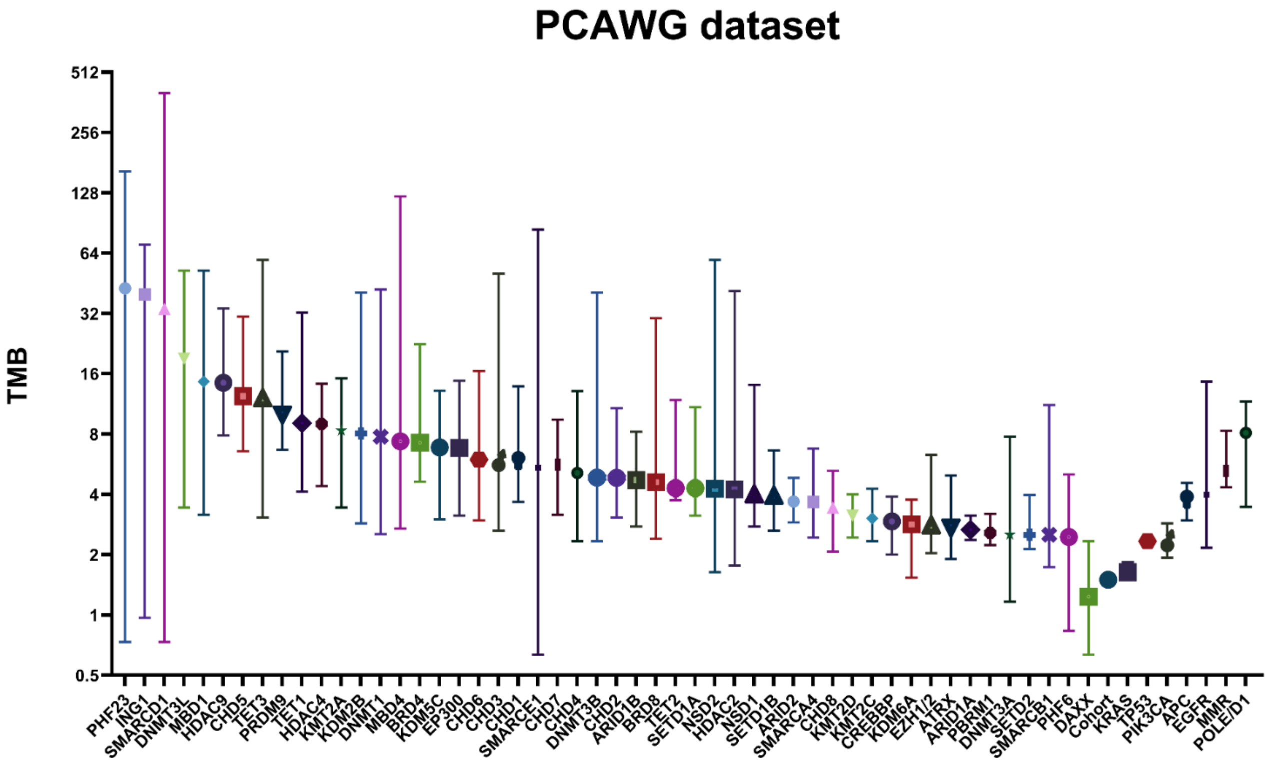


**Supplementary Figure 4**. Samples with mutations in single chromatin regulators (CRs), samples with mutations in canonical oncogenes and tumor-suppressor genes, and samples with mutations in MMR genes and *POLE* and *POLD1* were taken from the PCAWG dataset. The graph represents the median with 95% confidence intervals (CI). This figure is related to Figure 1. This figure is related to the section “Mutations in chromatin regulators are pervasive in cancer and associated with high tumor mutational burden”. The median of the analyzed samples is 49. The number of samples per subgroup used in this study can be found in Suppl. Data 9.

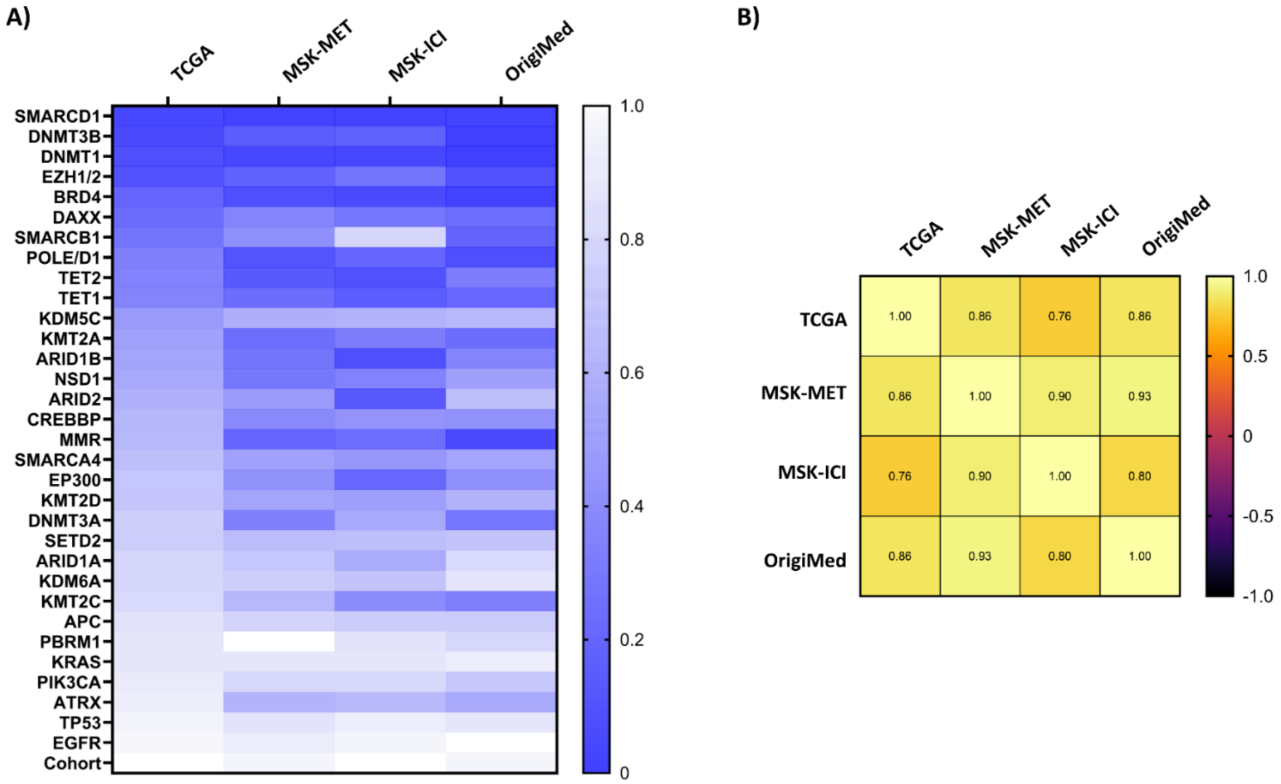


**Supplementary Figure 5**. The types of CRs associated with high TMB were reproducible across datasets. CRs were ranked in every dataset based on the strength of association with TMB. A) Heatmap of CRs and controls across datasets ranked based on the strength of their association with TMB, with 1 showing the weakest, while 0 showing the strongest association with TMB. The sorting is done according to the TCGA dataset. B) Heatmap of Pearson correlation coefficients, showing a high correlation of TMB-associated CRs between datasets. This figure is related to Figure 1. This figure is related to the section "Mutations in chromatin regulators are pervasive in cancer and associated with high tumor mutational burden".

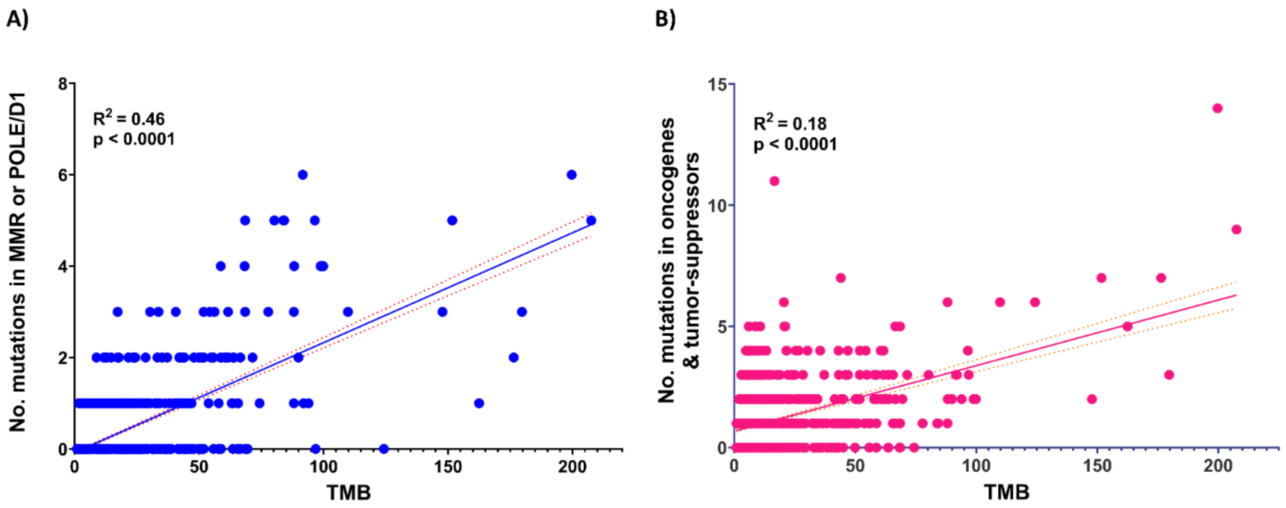


**Supplementary Figure 6**. Simple linear regression model showing that TMB cannot be equally well predicted based on the number of co-occurring mutations in multiple A) MMR and/or *POLE*, *POLD1* genes, B) traditional oncogenes and tumor suppressor genes in comparison to co-occurring mutations in CRs (Figure 2B). The datasets are taken from the MSK-ICI study. This figure is related to Figure 2. The figure is related to the section "The co-occurrence of mutations in chromatin regulators leads to additional increases in TMB."

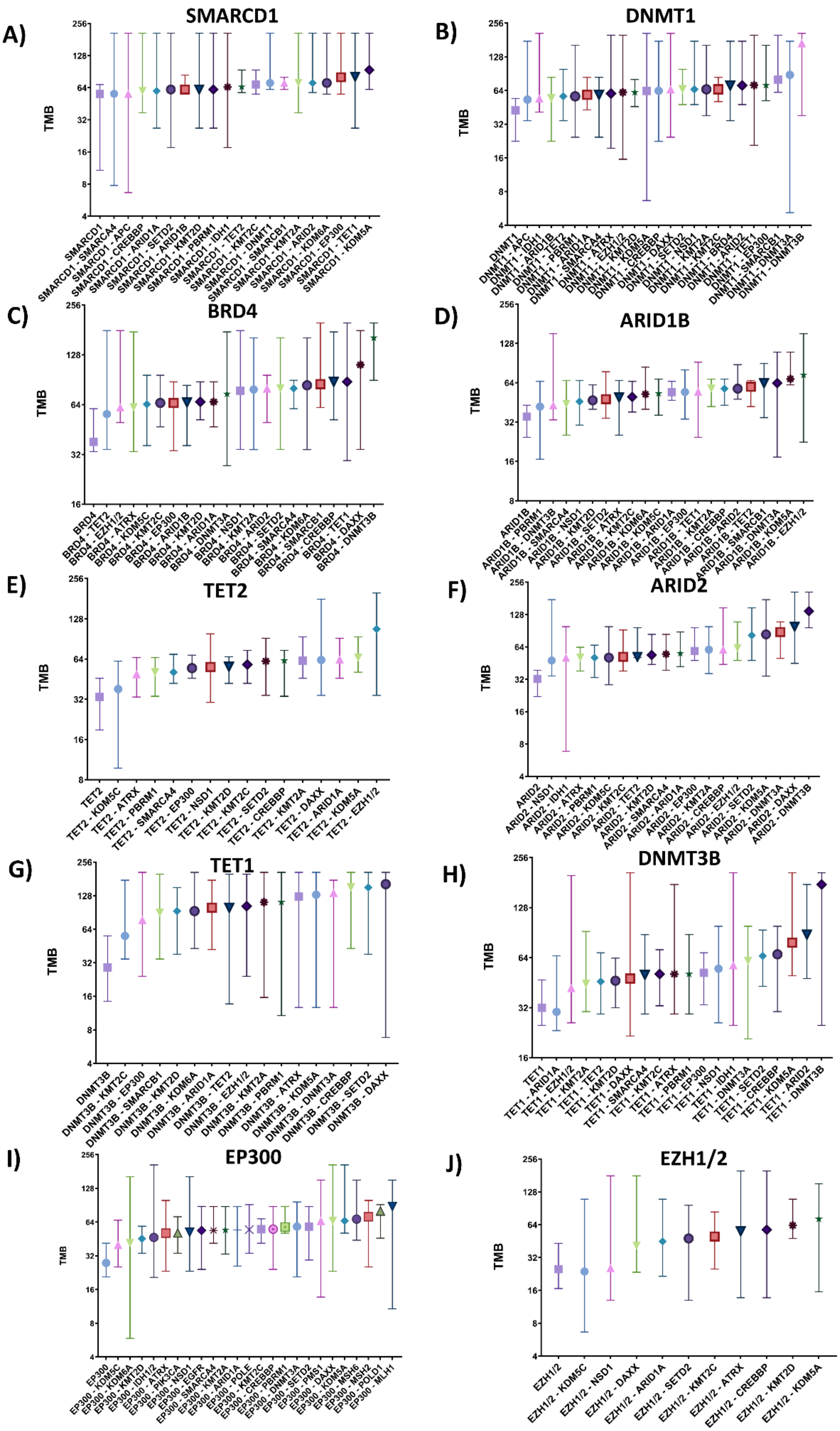
A)
SMARCD1
B)
DNMT1
C)
BRD4
D)
ARID1B
E)
TET2
F)
ARID2
G)
TET1
H)
DNMT3B
I)
EP300
J)
EZH1/2
TMB

**Supplementary Figure 7**. Co-occurrence of mutations in chromatin regulators leads to increased TMB. Statistically significant associations of co-occurrence between A) *SMARCD1*, B) *DNMT1*, C) *BRD4*, D) *ARID1B*, E) *TET2*, F) *ARID2*, G) *TET1*, H) *DNMT3B*, I) *EP300*, J) *EZH1* and *EZH2* with other CRs lead to further increase in TMB. The graph represents the median with 95% confidence intervals (CI). Datasets were taken from the MSK-ICI study. This figure is related to Figure 2. The figure is related to the section "The co-occurrence of mutations in chromatin regulators leads to additional increases in TMB". The number of samples per subgroup used in this study can be found in Suppl. Data 9.

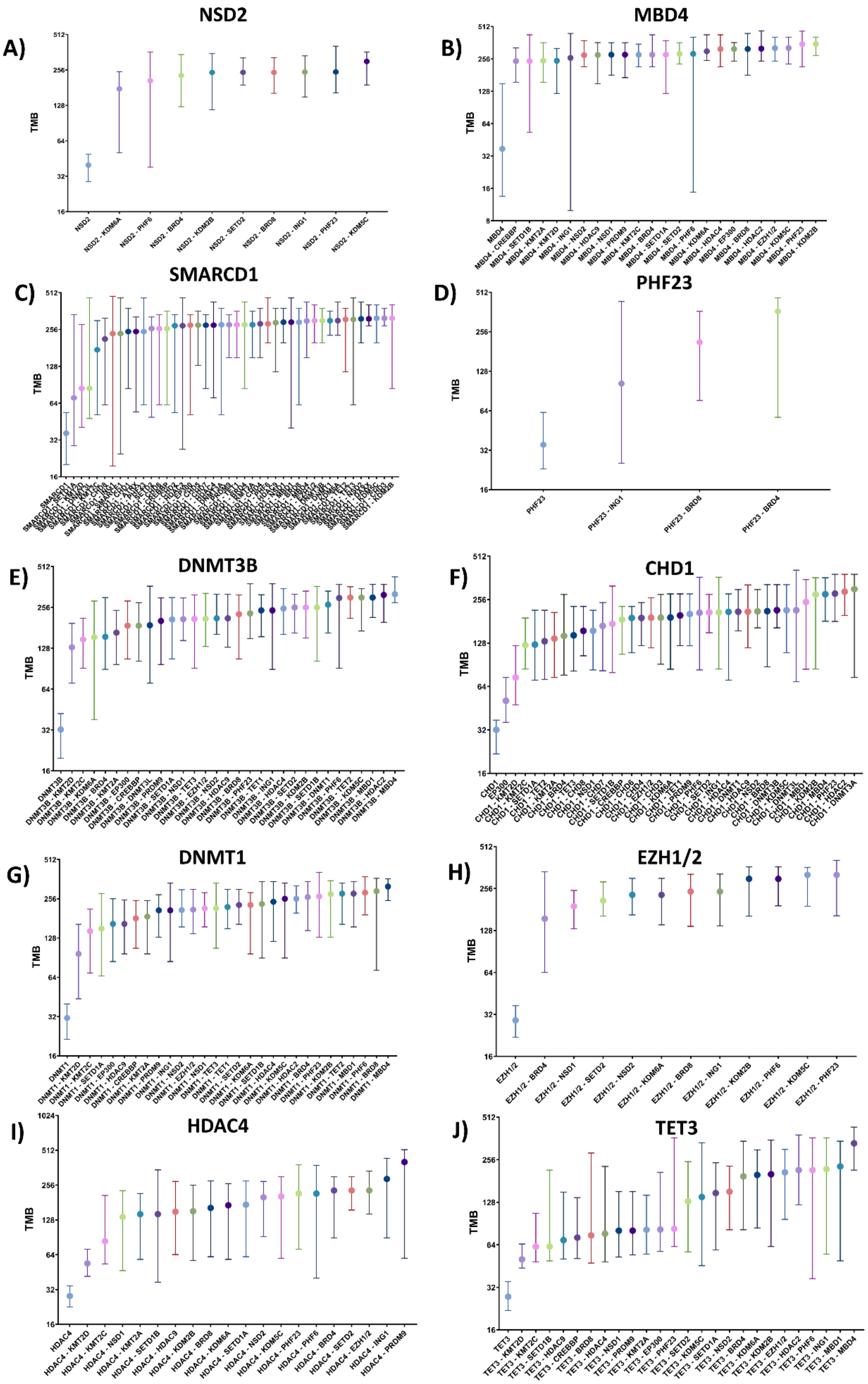
A)
NSD2
TMB
B)
MBD4
TMB
C)
SMARCD1
TMB
D)
PHF23
TMB
E)
DNMT3B
TMB
F)
CHD1
TMB
G)
DNMT1
TMB
H)
EZH1/2
TMB
I)
HDAC4
TMB
J)
TET3
TMB

**Supplementary Figure 8**. Co-occurrence of mutations in chromatin regulators leads to increased TMB. Statistically significant associations of co-occurrence between A) *NSD2*, B) *MBD4*, C) *SMARCD1*, D) *PHF23*, E) *DNMT3B*, F) *CHD1*, G) *DNMT1*, H) *EZH1* and *EZH2*, I) *HDAC4*, J) *TET3* with other CRs lead to further increase in TMB. The graph represents the median with 95% confidence intervals (CI). Datasets were taken from the TCGA study. This figure is related to Figure 2. The figure is related to the section “The co-occurrence of mutations in chromatin regulators leads to additional increases in TMB”. The number of samples per subgroup used in this study can be found in Suppl. Data 9.

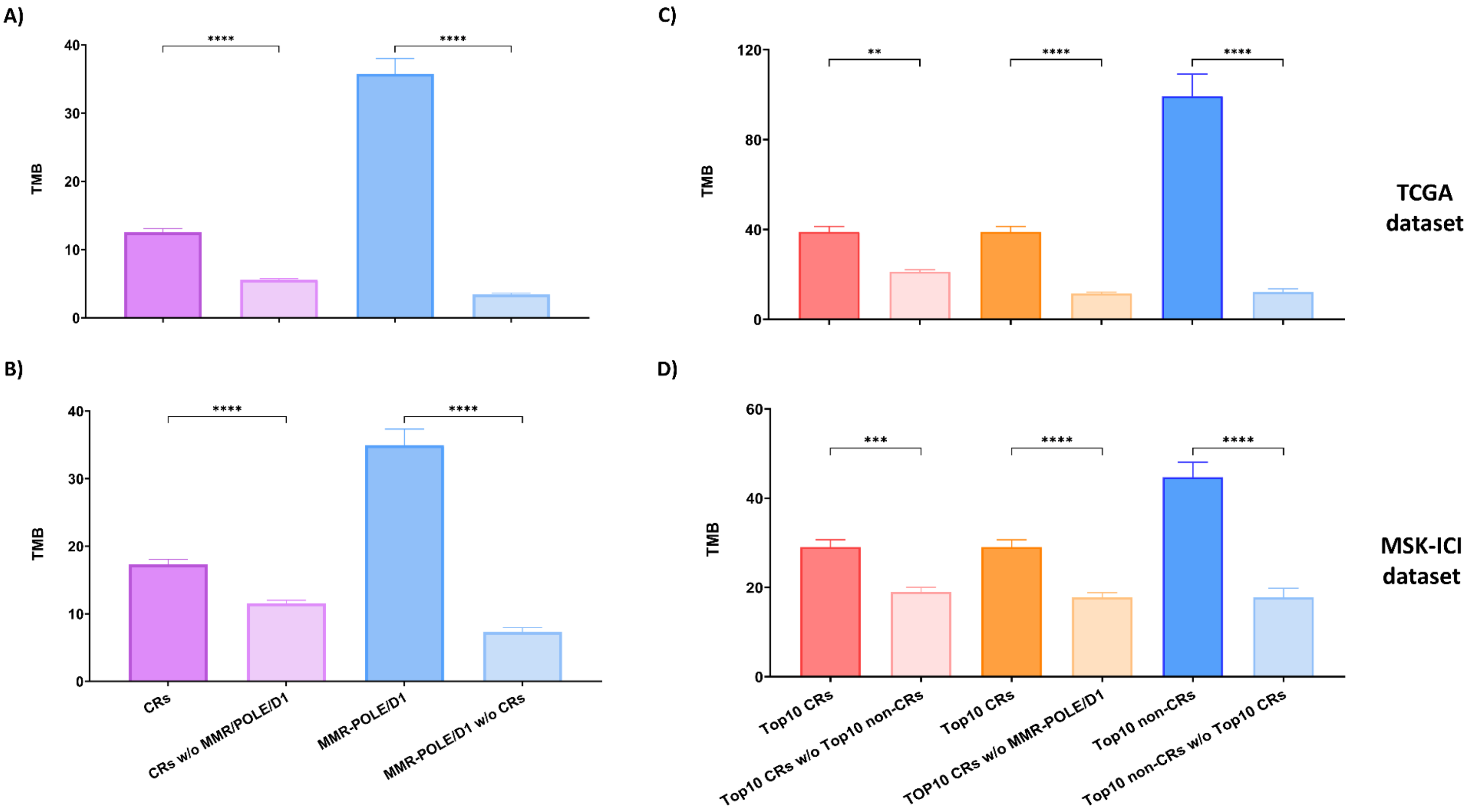
A)
B)
C)
D)
TMB
****
***
**
CRs
CRs w/o MMR/POLE/D1
MMR-POLE/D1
MMR-POLE/D1 w/o CRs
Top10 CRs
Top10 CRs w/o Top10 non-CRs
TOP10 CRs w/o MMR-POLE/D1
Top10 non-CRs
Top10 non-CRs w/o Top10 CRs
TCGA dataset
MSK-ICI dataset

**Supplementary Figure 9**. Co-occurrence and interdependence of mutations in chromatin regulators, and mutations in MMR genes or POLE/D1 genes in different patient subgroups and their relationship with TMB. A) Samples harboring mutations in the repair machinery MMR and/or POLE/D1 (n=1209), samples harboring mutations in the repair machinery MMR and/or POLE/D1 without co-occurring mutations in CRs (n=210), samples harboring mutations in CRs (n=5321), samples harboring mutations in CRs without co-occurring mutations in the repair machinery (MMR and/or POLE/D1) (n=4322), and their effect on TMB in the TCGA dataset. B) Samples harboring mutations in the repair machinery MMR and/or POLE/D1 (n=224), samples harboring mutations in the repair machinery MMR and/or POLE/D1 without co-occurring mutations in CRs (n=32), samples harboring mutations in CRs (n=935), samples harboring mutations in CRs without co-occurring mutations in the repair machinery (MMR and/or POLE/D1) (n=743), and their effect on TMB in the MSK-ICI dataset. C) Samples harboring mutations in TOP10 CRs (n=1091), samples harboring mutations in TOP10 CRs without co-occurring mutations in the repair machinery (MMR and/or POLE/D1) (n=651), samples harboring mutations in TOP10 CRs without co-occurring mutations in the TOP10 non-CRs (n=961), samples harboring mutations in TOP10 non-CRs (n=235), samples harboring mutations in TOP10 non-CRs without co-occurring mutations in the TOP10 CRs (n=107) and their effect on TMB in the TCGA dataset. D) Samples harboring mutations in TOP10 CRs (n=372), samples harboring mutations in TOP10 CRs without co-occurring mutations in the repair machinery (MMR and/or POLE/D1) (n=248), samples harboring mutations in TOP10 CRs without co-occurring mutations in the TOP10 non-CRs (n=280), samples harboring mutations in TOP10 non-CRs (n=143), samples harboring mutations in TOP10 non-CRs without co-occurring mutations in the TOP10 CRs (n=51) and their effect on TMB in the MSK-ICI dataset. The bars show the mean of the group, while the error bars represent the standard error of the mean (SEM). Statistical significance was calculated with the Dunn's multiple comparisons test. This figure is related to Figure 2. The figure is related to the section "The co-occurrence of mutations in chromatin regulators leads to additional increases in TMB". The number of samples per subgroup used in this study can be found in Suppl. Data 9.

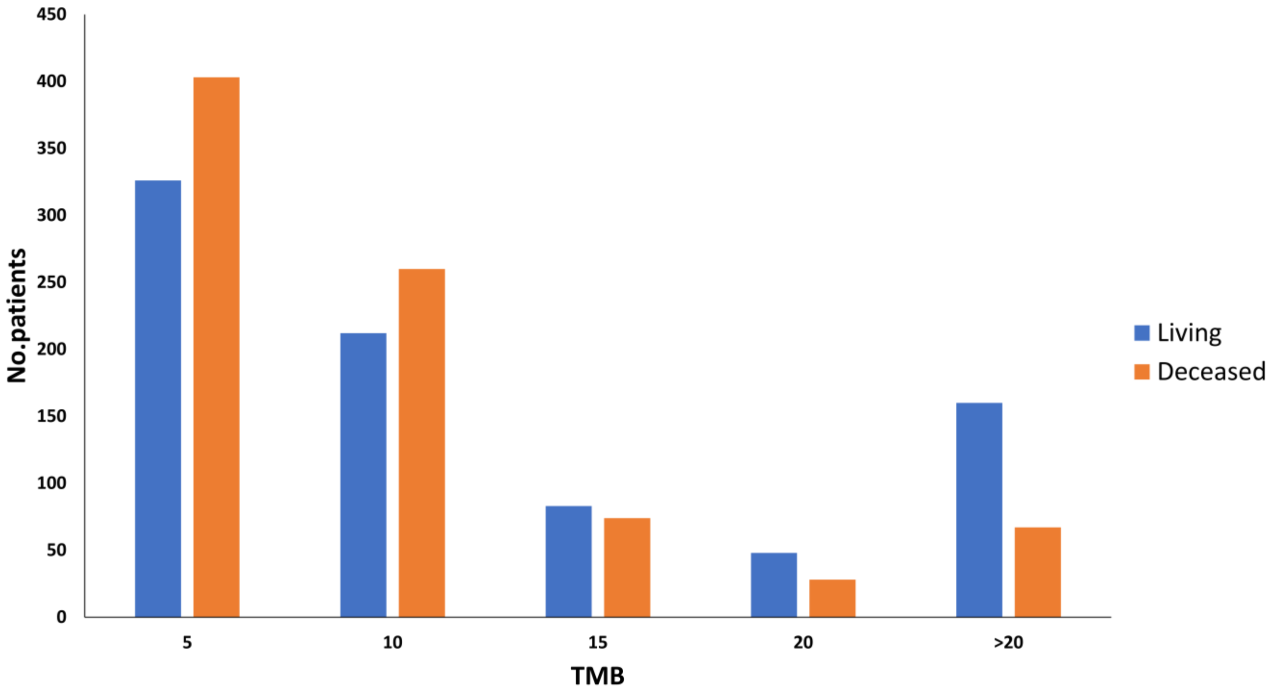


**Supplementary Figure 10**. The distribution of TMB levels in living and deceased ICI-treated patients. Datasets were taken from the MSK-ICI study. This figure is related to Figure 3.

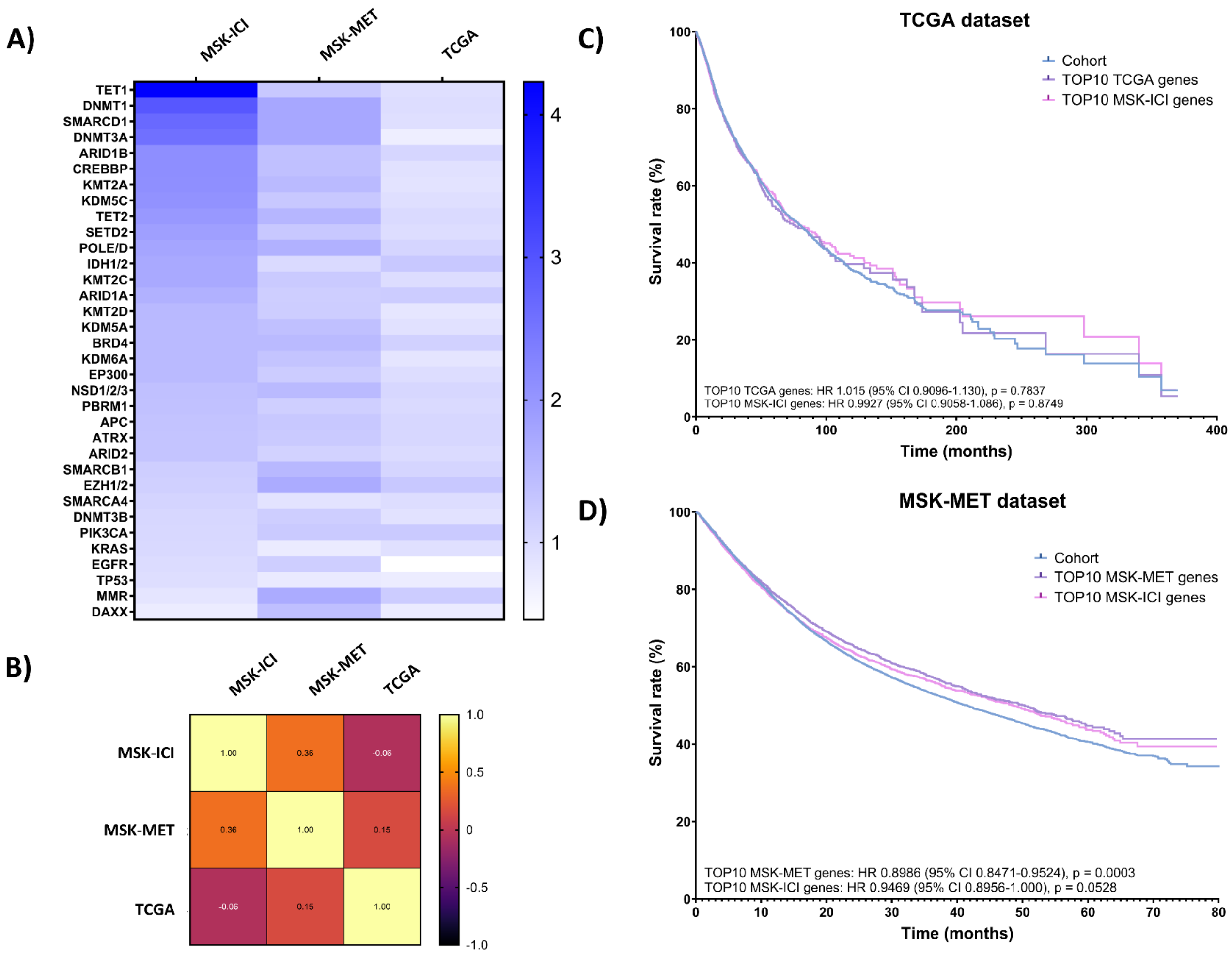


**Supplementary Data 11**. Survival after ICI treatment is dependent on the presence of mutations in CRs. A) Heatmap of the ratios of each CR in living and deceased patients in the ICI-treated group, and variously-treated groups (MSK-MET, TCGA), B) Heatmap of the Pearson correlation coefficient between the ratios of each CR in living and deceased patients in the ICI-treated group, and variously-treated groups (MSK-MET, TCGA), C) Kaplan-Meier survival analysis between patients with mutations in the TOP 10 high TMB CRs from the MSK-ICI cohort, TOP 10 high TMB CRs from the TCGA cohort compared with the TCGA cohort, D) Kaplan-Meier survival analysis between patients with mutations in the TOP 10 high TMB CRs from the MSK-ICI cohort, TOP 10 high TMB CRs from the MSK-MET cohort compared with the MSK-MET cohort. The Cox proportional hazards model was used to calculate hazard ratios. This figure is related to Figure 3. The figure is related to the section "Mutations in chromatin regulators lead to improved response to checkpoint immunotherapy".
